# Exploring the limits of LLMs in low-resource information extraction: Case study in brain MRI reports for Epilepsy

**DOI:** 10.1101/2025.08.02.25332570

**Authors:** Thinh Hung Truong, Emma Foster, Timothy Fazio, Sarah Holper, Karin Verspoor

**Affiliations:** University of Melbourne, Melbourne, Australia; RMIT University, Melbourne, Australia; Royal Melbourne Hospital, Melbourne, Australia; Alfred Health, Melbourne, Australia; Monash University, Melbourne, Australia

**Keywords:** information extraction, low-resource, LLM

## Abstract

Information extraction (IE) from specialized clinical texts such as brain MRI reports is important for various clinical and population health contexts. However, this topic is under-explored due to privacy concerns limiting data availability and the inherent complexity and domain-specificity of clinical language. Common methods relying on substantial amounts of training data fail. The recent advances in large language model (LLM) research provide a promising solution to bridge the data scarcity gap, with improved ability to adapt to novel tasks with little supervision. We introduce a new, challenging dataset of 100 expert-annotated brain MRI reports, featuring 152 fine-grained entity types and 4 relation types, characterised by low inter-annotator agreement. This task reflects the inherent complexity and real-world ambiguity of medical text.

We evaluate a small, open-weight LLM across span detection, named entity recognition, and relation extraction tasks. We compare few-shot prompting and parameter-efficient fine-tuning against specialized off-the-shelf biomedical IE systems. Our results demonstrate that both few-shot and fine-tuned LLM approaches substantially outperform off-the-shelf baselines. While LLMs show superiority, absolute performance, particularly for complex relations and fine-grained entities, remains modest, correlating with the dataset’s inherent difficulty and the extreme low-resource setting.

## 1. Introduction

Information extraction (IE) is a classic Natural Language Processing (NLP) problem, involving extraction of structured knowledge from free-text documents. IE encompasses different tasks such as extracting relevant spans from texts and classifying them into meaningful semantic categories (named entity recognition, NER), and identifying relations between entities (relation extraction, RE). IE is a well-explored task with many established benchmarks and methods.^1,2^ Despite significant progress, the most prevalent challenge for IE lies in its open-endedness: how to build a “universal” system capable of adapting to new domains, entity types, and relations with minimal effort. Most established methods for IE rely on a substantial amount of labeled training data to train supervised machine learning models.^3,4^ This reliance on large datasets represents a significant bottleneck for many specialized domains, such as clinical medicine, where annotated data is scarce, expensive to create, and governed by strict privacy regulations, while the set of relevant concepts to recognize is large. As a result, dictionary-based methods have often been preferred for concept recognition in these contexts.^5–7^

With the introduction of Large Language Models (LLMs), new possibilities have emerged to tackle these challenges, particularly leveraging their remarkable potential to perform tasks with minimal or no task-specific training data (zero-shot or few-shot learning).^8,9^ These models, pre-trained on vast amounts of text, often capture rich semantic knowledge that can be adapted for downstream tasks like IE through prompting or minimal fine-tuning.

In this work, we explore the feasibility of using LLMs for low-resource IE in the highly specialized domain of radiology. As a case study, we utilize brain MRI reports annotated with clinical concepts and relations relevant for Epilepsy. With only 100 annotated reports, encompassing a rich set of 152 entity types and 4 relation types, we investigate the capabilities and limitations of LLMs in achieving meaningful extraction performance. We break down the complex IE problem into distinct task formulations based on the granularity of extraction required: span-only identification, entity types, and relation extraction. Each task formulation is evaluated using both standard fine-tuned models (as baselines where applicable) and various approaches leveraging LLMs. We experiment with different settings for applying LLMs: zero-shot prompting, few-shot prompting (providing examples in the prompt), and fine-tuning. We explore both end-to-end generation with a single prompt encompassing all tasks, and pipeline approaches where the output from one task (e.g., entity extraction) informs the prediction for the next (e.g., relation extraction).

In general, we make two main contributions:

- We introduce a new, challenging dataset for information extraction from brain MRI reports. It features fine-grained annotations covering a diverse set of Epilepsy-related clinical entity types and their relationships, reflecting real-world complexity.
- We explore the practical limits of applying a small, open-weight LLM (capable of running on consumer-grade GPUs) to this task. This demonstrates a realistic scenario for institutions where data privacy is paramount and reliance on external, large-scale proprietary models is not feasible.

## 2. Related work

In traditional NLP, Information Extraction (IE) is typically framed as a sequence labeling task. Methods often involve fine-tuning pre-trained encoder models, such as BERT^10^ or its domain-specific variants (e.g., BioBERT^3^ or clinical BERT models^11^), with an additional classification head (e.g., a linear layer) to predict a label for each token using the BIO tagging scheme.^12^ These approaches have achieved state-of-the-art results on many benchmarks when sufficient labeled data is available.^4^ In contrast, using generative methods based on LLMs, IE is framed as a text-to-text task. The model takes the source text as input and is prompted to generate the relevant text spans along with their corresponding labels or structures directly, often in a structured format like JSON, or as natural language statements.^13^

LLMs have been explored for IE tasks with promising results.^14^ While early studies suggested that LLMs might lag behind smaller, fine-tuned encoder models on classic benchmarks with ample supervision signals,^15^ their strength lies in adaptability and performance in low-resource or zero-shot settings. Generative methods are gaining traction due to their ability to perform well in new domains or on novel entity/relation types by simply modifying the prompt or providing a few relevant examples (in-context learning).^8^ Specifically for biomedical text, LLMs have been applied to extract medical problems, tests, and treatments from patient discharge summaries.^16^ The authors showed that using only 8 few-shot samples and carefully curated prompts, LLMs could achieve comparable performance to fully-supervised systems on the popular i2b2 dataset.^17^ However, the setting is relatively simple with only 3 entity types and 8 relations and a substantial number of training samples. Radiology reports, especially highly structured ones like brain MRIs, present unique challenges. They combine structured or semi-structured templates with free-text descriptions, utilize highly specific anatomical and pathological terminology, and often contain implicit relations that require nuanced understanding. Furthermore, the clinical requirement for high precision makes IE particularly demanding in this domain.

Different strategies for prompting LLMs for IE have been explored, ranging from simple instruction-based prompts to more complex schemes involving structured output formats,^18^ task reformulation,^19^ or chain-of-thought reasoning.^20^ While extremely large models demonstrate impressive capabilities, recent research increasingly focuses on the potential of smaller, open-weight models (e.g., Llama,^21^ Flan-T5^22^) for domain-specific tasks.^23^ These models offer a pathway for deploying powerful NLP capabilities locally in secure environments, mitigating data privacy concerns and reducing computational costs, which is highly relevant for medical applications. Our work adds to this research by systematically evaluating a smaller LLM’s effectiveness on a complex, low-resource medical IE task through both prompting and fine-tuning.

## 3. Dataset: Brain MRI reports

Brain MRI reports from a cohort of epilepsy patients were obtained from The Royal Melbourne Hospital, a major comprehensive epilepsy centre in Melbourne, Australia. A semi-randomised approach was used to select the reports, to ensure they were sampled over a period of years and from different reporting radiologists to capture variations in phrasing, sentence structure, and reporting styles. The reports were deidentified by clinicians to remove dates, locations, names prior to annotation.

### 3.1. Annotation Schema

#### 3.1.1. List of entities

A hierarchical list of entities was collated and refined for the MRI annotation process. This list was based on terminology published by the National Institute of Neurological Disorders and Stroke (NINDS), Epilepsy / Imaging Diagnostics / MRI Common Data Elements^a^.^25^ The NINDS common data elements were first released in 2010 and were compiled by subject-specific working groups to promote consistency for naming and collecting data elements in epilepsy research. This initiative aims to improve efficiency and effectiveness of clinical research studies and clinical treatment, increase data quality and facilitate data sharing. Researchers also added MRI entities described in a real-world study of a well-characterised cohort of patients with new-onset seizures.^26^ The final list of entities adopted for this work is displayed in Appendices A-B.

### 3.2. Relations

Hedging and negation terms indicating the possible or negative presence of entities were also targeted. For instance, “Differentials include… “(*possible* hedging), “There is no evidence of… “(*negation*). These terms were then linked to the corresponding entities to capture the scope of the relation. Negation was grouped with terms suggesting a finding was unlikely. No hedging term was assigned when a high degree of certainty was given for the presence of an entity, e.g., “This most likely represents… “. In these instances, the entity alone was annotated.

Formally, 4 distinct relation types are defined:

- `unlikely-rel`: Indicates negation or low likelihood of a finding.
- `possible-rel`: Indicates uncertainty or possibility of a finding.
- `location-rel`: Links a finding (lesion, feature) to its anatomical location.
- `feature-rel`: Links an entity (e.g., a lesion) to its descriptive features (e.g., size, signal characteristics).

### 3.3. Annotation process

Following refinement of the list of entities and hedging terms, two board-certified neurologists annotated the reports according to the schema. An example appears in Table 1.

**Table 1.**
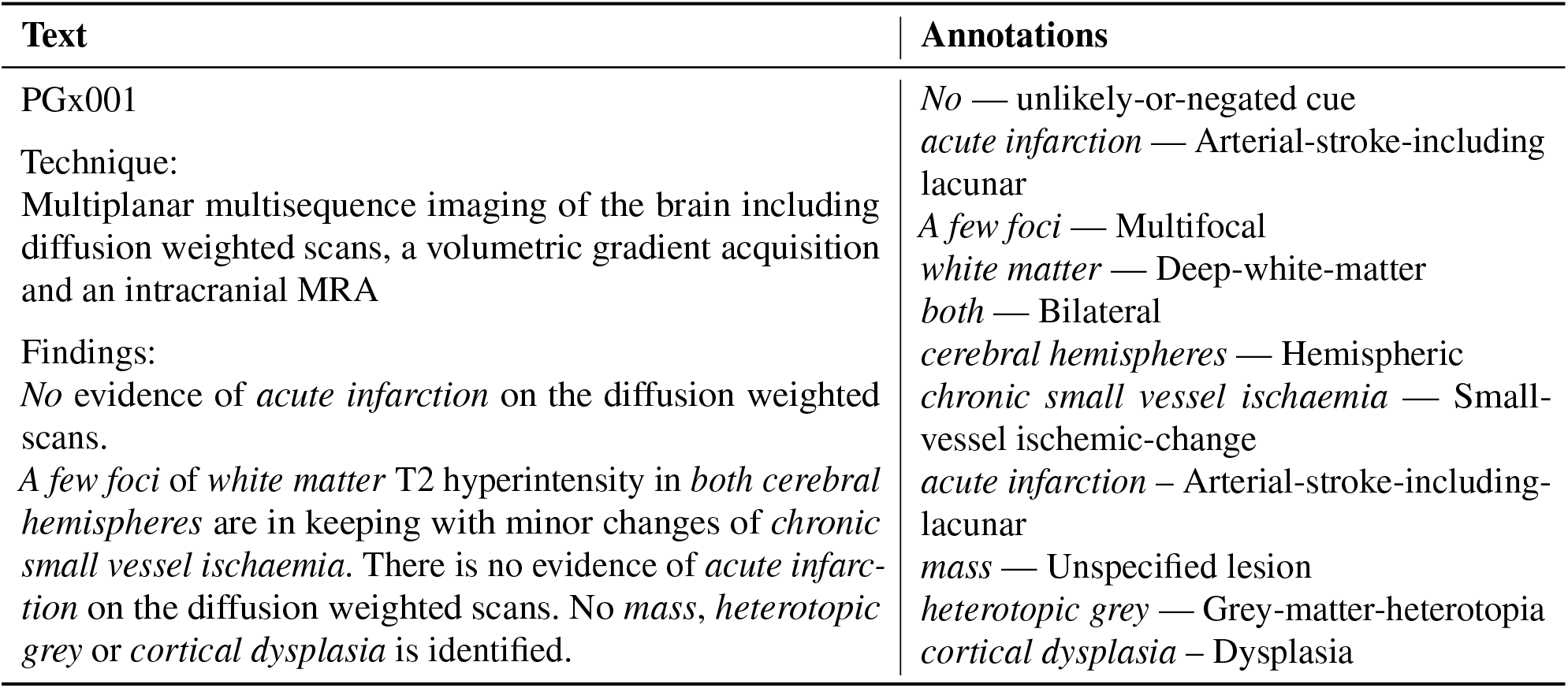
Extract from a radiology report with selected entity annotations, converted from brat^24^ standoff format for clarity.

| Text | Annotations |
| --- | --- |
| PGx001 | *No* — unlikely-or-negated cue |
| Technique: | *acute infarction* — Arterial-stroke-including lacunar |
| Multiplanar multisequence imaging of the brain including diffusion weighted scans, a volumetric gradient acquisition and an intracranial MRA | *A few foci* — Multifocal |
|  | *white matter* — Deep-white-matter |
|  | *both* — Bilateral |
| Findings: | *cerebral hemispheres* — Hemispheric |
| *No* evidence of *acute infarction* on the diffusion weighted scans. | *chronic small vessel ischaemia* — Small-vessel ischemic-change |
| *A few foci* of *white matter* T2 hyperintensity in *both cerebral hemispheres* are in keeping with minor changes of *chronic small vessel ischaemia*. There is no evidence of *acute infarction* on the diffusion weighted scans. No *mass*, *heterotopic grey* or *cortical dysplasia* is identified. | *acute infarction* – Arterial-stroke-including-lacunar |
|  | *mass* — Unspecified lesion |
|  | *heterotopic grey* — Grey-matter-heterotopia |
|  | *cortical dysplasia* – Dysplasia |

#### 3.3.1. Annotation environment

The de-identified MRI reports were uploaded into brat,^24^ an open-source web-based annotation tool, installed on a hospital firewall- and password-protected server. Clinician annotators highlighted words or phrases in the text, and assigned one or more entities from a drop-down list. For example, “stroke” was annotated as “arterial stroke”, “oligodendroglioma” was annotated as “tumour / primary”, etc.

#### 3.3.2. Iterative annotation

The clinicians co-annotated the first 10 MRI reports together to gain an understanding of the annotation process and agree on a mutual approach. Subsequent MRI reports were annotated independently, in batches of 10, followed by assessment of inter-annotator agreement (IAA), based on treating one neurologist as the “gold standard”. Annotation differences between reports were marked as “false negative” and “false positive”, referring to either missed or additional annotations with reference to the gold standard. Differences in annotation spans and linking of hedging terms were also noted. These anotations were merged on the MRI reports in brat, and reviewed together by the neurologists. Differences were resolved by consensus. An example of annotation variation is the phrase “ventricular enlargement”, which could be annotated as both “atrophy” and “ventriculomegaly”, depending on the context provided in the remainder of the report. Once neurologists reached acceptable levels of inter-annotator agreement, which was achieved after approximately 50 MRI reports, they proceeded to annotate separate MRI reports to increase the number ultimately included in the training set.

### 3.4. Dataset statistics

The details of the dataset are provided in Table 2. In total, there are 101 reports annotated with 102 different entity types and 4 relations. We calculate the initial inter-annotator agreement between two annotators using F1-score. The F1-score for the entity-level annotation is just around 60% with 25 labels with 0 agreements, and relation-level annotation has below 10% agreement, showing that this is a hard task even for human experts.

**Table 2.** Distribution of Annotations Across Levels with Inter-Annotator Agreement and Unique Counts.

| Level | Count | Unique | IAA ($F_1$) |
| --- | --- | --- | --- |
| Spans | 2558 | 790 | 70.3% |
| Entities |  |  |  |
| Entities | 1274 | 586 | 47.7% |
| Location | 591 | 107 | 70.0% |
| Other | 149 | 69 | 28.9% |
| Hedging/Negation | 544 | 95 | 83.3% |
| Relations |  |  |  |
| unlikely-rel | 703 | 374 | 8.3% |
| location-rel | 81 | 72 | 3.8% |
| possible-rel | 62 | 60 | 0.0% |
| feature-rel | 34 | 32 | 0.0% |

Overall, we find that higher diversity leads to lower IAA. Unsurprisingly, relation annotation was the most difficult, as the agreement need to be reached for 3 elements — the head and tail entities, and the relation between them. In addition to having high diversity, the relation distribution also skews heavily toward `unlikely-rel`. Both `possible-rel` and `feature-rel` have the highest diversity while having 0% IAA, showing severe unreliability. We treat IAA as the human-level upper-bound performance. The low IAA across all levels display the difficulty of this task.

## 4. Experiments

### 4.1. Task formulation

Following previous works in generative IE, we formulate this task as a text-to-text problem where the corresponding list of entities or relation triples are generated from raw input text. This is different from traditional sequence labeling tasks, where each token is classified as being inside or outside an entity or relation span.

The annotations are captured in the brat standoff format,^24^ which includes precise character offsets for each annotation. Based on the level of granularity required for evaluation and to better suit the generative text-to-text task, we convert the brat annotation file into a scheme focused on the text span and its label, omitting the explicit start and end offsets. This conversion simplifies the target output for the generative models, which struggle to produce precise character indices. This process may result in identical text spans annotated with the same label at different positions being treated as a single instance in the target output or indeed overgenerations;^27^ we removed such duplicates from the target list for simplicity during training and evaluation of the models.

#### Clinical concept span detection

As the simplest task formulation, we focus solely on identifying spans of relevant clinical concepts within the report, without assigning any specific entity or relation type. The target output is simply a list of extracted text spans corresponding to any annotated concept. This assesses the model’s fundamental ability to locate salient information snippets.

#### Named entity recognition

For named entity recognition, the goal is to extract text spans and classify them with their corresponding entity type. Each target annotation is formatted as `extracted entity span - entity type.` The annotation scheme used (Appendices A-B) is hierarchical, with up to 4 levels of specificity; an entity label may belong to any of these levels.

In this work, we evaluate NER performance at two primary levels of granularity to understand the model’s capabilities at different abstraction levels:

- **Coarse:** Using only the outermost category label (e.g., Vascular-malformation).
- **Fine:** Using the most specific sub-category label available (e.g., Arteriovenous-malformation, Cavernous-angioma, Cerebral-aneurysm, Developmental-venous-anomaly, which are all sub-types of Vascular-malformation).

Evaluating at both the Coarse and Fine levels allows us to assess the model’s ability to capture broader semantic categories versus more specific, granular concepts present in the text.

#### Relation extraction

For relation extraction, the task is to identify relationships between pairs of extracted entities. Each target annotation is a triple formatted as: `head entity span - relation type - tail entity span.`

To improve the quality of generated relations and reduce invalid outputs, we explicitly define the permissible entity types for the head and tail arguments of each relation type, information also available during manual annotation via constraints specified in the brat configuration. These constraints are also incorporated into the LLM prompt instructions (see Appendix D) to guide the generation process. For example, the type constraints for the `location-rel` relation are described as in Figure 1. An overall summary of the relations adopted appears in Appendix C.

**Fig. 1.**
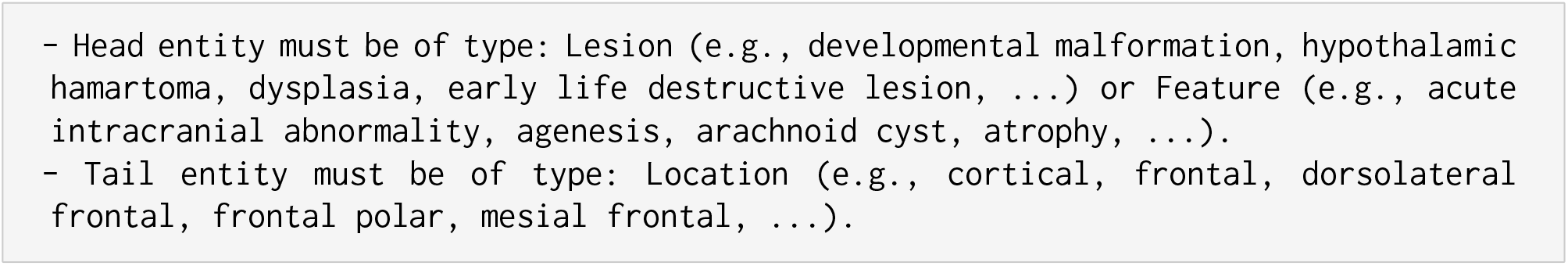
Constraint Example (excerpt) for location-rel. Note that the full list of eligible sub-types provided in the actual prompt is truncated here for clarity.

### 4.2. Baselines

For all the above settings, we consider different baselines that can be applied to this type of task:

#### Dictionary-based

We perform a straightforward dictionary lookup using the label spaces provided in the annotation schemes, with some simple text normalization steps (e.g. removing hyphen, lower-casing). For instance, for the Entity-level extraction, the label `Grey-matter-heterotopia` will be normalized into “grey matter heterotopia” for exact matching in the text. If such a span exists, we will tag it with the same label (Grey-matter-heterotopia). For Relation-level, we group the 3 closest eligible matches of head entity, relation, and tail entity to form a triple.

#### Off-the-shelf system

We use SciSpacy,^28^ a system for biomedical information extraction. For entity-level, we employ a retriever based on to retrieve the top-1 match from the extracted entities from SciSpacy and the label set from the annotation scheme. For relation-level, we employ the CueNB^29^ for `unlikely-rel` and NegEx^30^ for `possible-rel`, while for `location-rel` and `feature-rel,` we use OpenIE employed in Stanza^31^ and map is-in relations to `location-rel,` and has- or is-relations to `feature-rel.`

#### Few-shot

This represents our main approach. We utilize a powerful, open-weight LLM that can be run locally on consumer-grade hardware, ensuring data privacy and demonstrating practical applicability. Specifically, we use Llama-3-8B-Instruct.^21^ We employ few-shot prompting, where the prompt includes instructions defining the task and output format, followed by a small number (k=10) of randomly selected examples (demonstrations) from our annotated dataset. We found that using 10 samples provided a reasonable balance between context length and performance; preliminary experiments suggested that adding significantly more samples yielded only small marginal improvements for this specific model and task setup.

The general structure of the prompt is shown in Figure 2. The format of each {annotation} line within the examples depends on the specific task formulation:

**Fig. 2.**
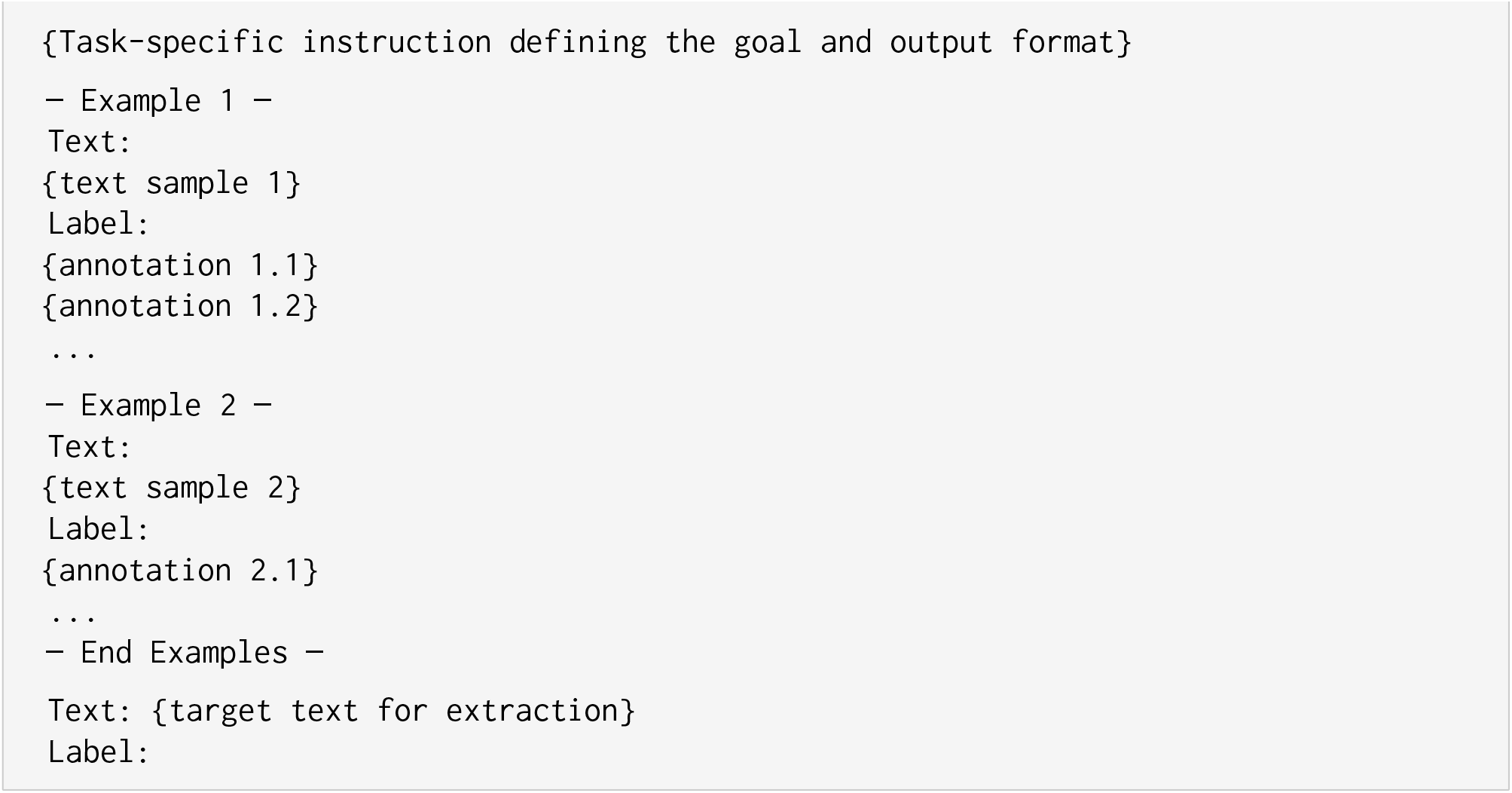
Template for a few-shot prompt.

1. Relevant Span detection: Annotation is the text span, e.g., *mild diffuse cerebral volume loss*.
2. NER: Annotation includes the span and type, e.g., *mild diffuse cerebral volume loss – Atrophy*.
3. RE: Annotation is the relation triple, e.g., *mild diffuse cerebral volume loss –* location-rel*– cerebral*.

The *Task-specific Instruction* also varies for each task (Span, Entity, Relation) and includes details like the entity type constraints for RE. For NER and RE, we also include the list of valid labels in the instruction. The full list of entity types and constraints for relation extraction are in the Appendices.

As a post-processing step, particularly for NER and RE where the LLM might generate entity types or text spans that don’t perfectly align with the gold standard annotations or predefined ontology,^27^ we employ a grounding strategy. This maps potentially out-of-vocabulary type labels generated by the LLM to the label options within our annotation scheme that are semantically closest, measured using cosine similarity of sentence embeddings derived from a pre-trained model. In particular, we used the BioLORD model,^32^ which has state-of-the-art performance on biomedical and clinical semantic similarity tasks. Similarly, generated entity spans can be loosely matched to ground truth spans if needed for evaluation metrics that allow for fuzzy matching.

#### Fine-tuning

We perform parameter-efficient fine-tuning using LoRa^33^ to fine-tune a LLama 3.1 8B model with MRI data converted into Alpaca instruction-tuning format.^34^ Fine-tuning details are in Appendix E. The models are evaluated following a 5-fold cross-validation with 20/80 train/test ratio. We keep the test set size larger to reflect the real-world scenario where ground truth data are scarce, and to make this comparable with the few-shot setting.

### 4.3. Main results

The main results for all 3 task formulations using the 3 baselines are in Table 3. For all settings, we adopt the standard Precision, Recall, F1-score metrics for information extraction tasks. We utilize the strict evaluation setting for relation extraction, where we consider a prediction to be correct only if the all 3 of the head entity, the tail entity, and the relation type are correct.

**Table 3.** Performance of the tested baselines for each level. Results are reported as mean (±std) over 5 random seeds. Span-level does not have Pipeline results as it is the first step in the pipeline.

| Task Level | Metric | Dictionary | Off-the-shelf | Few(10)-shot | Pipeline | Finetuning |
| --- | --- | --- | --- | --- | --- | --- |
| Span | F1 | 0.197 ($\pm 0.007$) | 0.212 ($\pm 0.007$) | 0.499 ($\pm 0.011$) | — | **0.598** ($\pm 0.020$) |
| | Precision | 0.532 ($\pm 0.018$) | 0.156 ($\pm 0.005$) | 0.550 ($\pm 0.017$) | — | **0.599** ($\pm 0.015$) |
| | Recall | 0.126 ($\pm 0.005$) | 0.352 ($\pm 0.012$) | 0.456 ($\pm 0.011$) | — | **0.597** ($\pm 0.029$) |
| Entity-Fine | F1 | 0.152 ($\pm 0.010$) | 0.110 ($\pm 0.009$) | 0.133 ($\pm 0.024$) | 0.140 ($\pm 0.041$) | **0.386** ($\pm 0.048$) |
| | Precision | **0.459** ($\pm 0.026$) | 0.077 ($\pm 0.007$) | 0.125 ($\pm 0.023$) | 0.115 ($\pm 0.038$) | 0.340 ($\pm 0.041$) |
| | Recall | 0.094 ($\pm 0.006$) | 0.205 ($\pm 0.011$) | 0.144 ($\pm 0.026$) | 0.172 ($\pm 0.047$) | **0.448** ($\pm 0.058$) |
| Entity-Coarse | F1 | 0.131 ($\pm 0.007$) | 0.177 ($\pm 0.011$) | 0.388 ($\pm 0.033$) | 0.375 ($\pm 0.015$) | **0.429** ($\pm 0.047$) |
| | Precision | **0.391** ($\pm 0.024$) | 0.129 ($\pm 0.009$) | 0.340 ($\pm 0.029$) | 0.324 ($\pm 0.014$) | 0.372 ($\pm 0.041$) |
| | Recall | 0.081 ($\pm 0.012$) | 0.303 ($\pm 0.004$) | 0.452 ($\pm 0.040$) | 0.465 ($\pm 0.043$) | **0.513** ($\pm 0.058$) |
| Relation | F1 | 0.000 ($\pm 0.000$) | 0.047 ($\pm 0.005$) | **0.222** ($\pm 0.019$) | 0.143 ($\pm 0.018$) | 0.195 ($\pm 0.029$) |
| | Precision | 0.000 ($\pm 0.000$) | 0.052 ($\pm 0.006$) | **0.207** ($\pm 0.018$) | 0.126 ($\pm 0.016$) | 0.127 ($\pm 0.020$) |
| | Recall | 0.000 ($\pm 0.000$) | 0.049 ($\pm 0.005$) | 0.240 ($\pm 0.021$) | 0.165 ($\pm 0.021$) | **0.423** ($\pm 0.064$) |

**Table 4.** Relation extraction results breakdown by type (F1 Scores)

| Relation Type | Few(10)-shot | Pipeline | Finetuning |
| --- | --- | --- | --- |
| unlikely-rel | 0.258 ($\pm 0.215$) | 0.179 ($\pm 0.192$) | **0.294** ($\pm 0.190$) |
| possible-rel | **0.042** ($\pm 0.131$) | 0.021 ($\pm 0.117$) | 0.016 ($\pm 0.092$) |
| location-rel | **0.037** ($\pm 0.146$) | 0.013 ($\pm 0.092$) | 0.016 ($\pm 0.069$) |
| feature-rel | **0.003** ($\pm 0.033$) | 0.002 ($\pm 0.028$) | 0.000 ($\pm 0.000$) |
| Overall | **0.222** ($\pm 0.017$) | 0.143 ($\pm 0.013$) | 0.195 ($\pm 0.029$) |

#### LLMs outperform specialized off-the-shelf systems

Across all task formulations, general-purpose large language models (LLMs) consistently outperform specialized off-the-shelf systems tailored for individual clinical or biomedical tasks, even with only a few demonstrations. Despite this, the absolute performance across all tasks remains relatively low. Only in the simplest span detection task do LLMs demonstrate relatively strong performance. Nevertheless, when using the initial inter-annotator agreement (IAA) score (Table 2) as a proxy for human performance, the LLMs’ output is not far behind. Moreover, LLMs offer a significant advantage in terms of ease of use and simplicity: they operate in an end-to-end fashion, unlike traditional systems that require multiple pipeline steps (e.g., extracting spans, assigning entity labels, and predicting inter-entity relations) or ample training data.

#### LLMs can learn task-specific patterns with sufficient supervision

While overall performance remains modest, we observe a promising trend in tasks with greater supervision—such as relevant span detection, named entity recognition, and the `unlikely-rel` relation—where LLMs perform noticeably better. In contrast, the poor performance on tasks like extracting location, feature and possible relation can be attributed to the high variability in the training data. Specifically, 32 out of 34 instances of `feature-rel` are unique, making it difficult for the model to generalise. The open-ended nature of this type of relation further complicates learning. Fine-tuning also yields larger gains in tasks where more training supervision is available (`unlikely-rel`, entities extraction). Overall, finetuning LLM achieved the best performance across most setting, outperforming other baselines by a large margin except for relation extraction. We also experimented with a pipeline formulation of the extraction task, wherein the output of one step is used as additional input in the prompt for the next step. For example, in entity-level extraction, we first identify entity spans, then condition the prompt on these spans to predict the corresponding entity types. While this structured approach provides intermediate supervision, it adds complexity compared to end-to-end approaches, and did not achieve any improvement.

#### Normalization is essential when applying LLM for IE tasks

When using LLMs for structured prediction, it is important to apply normalization to standardize the prediction, ensuring models don’t make up terms and concepts that are not in the target label spaces. In our experiments, we did find that normalization of spurious outputs was required with high frequency. In particular, the normalization rate is 36.8% for entities, and 20.1% for relations. This is not surprising given that we include a long context in the prompts (with the full list of labels, together with 10 examples of long input MRI reports-ground truth annotations). Another pervasive error is that the model generates predictions with invalid formats (e.g. truncated text spans, using the incorrect delimiters of the required, missing labels), produces entities that are not in the text,^27^ or hallucinates a new MRI report after generating. These were dismissed instead of normalizing.

### 4.4. Error analysis

We analyze the errors of the best performing setting (Fine-tuning) and categorise the most common errors in entity and relation extraction.

#### 4.4.1. Entities

Entity-level extraction suffers from over- or under-specificity. Models may label entities either too broadly or too narrowly. A clear example is seen in: “*Focal area of encephalomalacia, with surrounding* ***gliotic change*** *consistent with an old right parietal infarct*…” The model incorrectly assigns a generic category (`Post-traumatic-or-post-ischemic-lesion`) rather than the correct more specific sub-category general label (`Post-stroke`). This error can also goes in the other direction; it uses a more specific, yet incorrect label, when the label higher up in the hierarchy would be more accurate, e.g., *“*…***pituitary*** *gland measures 14mm tall, greater than expected for age* The model assigns a specific but inaccurate sub-category (Basal-ganglia), when the more generic label higher up in the hierarchy would be correct in this instance (`Subcortical`).

Additionally, models often confuse pathological processes with similar lexical forms. For example, they may mislabel *ischaemia* as `Small-vessel-ischemic-change` instead of the correct category `Arterial-stroke-including-lacunar`, due to surface-level lexical overlap. Finally, confusion between negation and possibility cues is a recurring issue, reflecting the subtlety and variability in clinical language, as well as the insensitivity of LLM on negation and hedging.^35^

#### 4.4.2. Relations

Across all relation types, incorrect entity boundaries remain the most dominant source of error. Consistent with prior NER research,^36^ issues related to span expansion and contraction are espe-cially prevalent. For example, the head entity “*enhancement*” is often misrecognized as “*abnormal enhancement*”, reflecting a common span inflation error.

This challenge also extends to negation cues, where variations in phrasing (e.g., “*no*” vs. “*no evidence*”) contribute to inconsistencies in model predictions. Radiology texts often feature complex noun phrases, such as “*hippocampi size, signal, and morphology*”, which further complicate entity recognition and relation extraction. Another significant source of error is the confusion between relation types, particularly `unlikely-rel` and `possible-rel`. These mistakes frequently arise from the subjective nature of radiological assessment, where radiologists may express varying degrees of diagnostic confidence.

The low performance on `location-rel` relations underscores the current models’ lack of spatial reasoning capability. This issue is exacerbated in radiology by the hierarchical structure of anatomical references, such as *hemisphere→lobe→gyrus*. Errors often stem from mismatches in anatomical specificity, such as predicting “*cortical*” when the correct span is “*left inferior frontal gyrus*”.

## 5. Conclusion

In this work, we investigated the capabilities of LLM for information extraction from brain MRI reports in an extreme low-resource setting. We introduced a novel, challenging dataset comprising 101 expert-annotated reports, characterised by its fine-grained annotation scheme (152 entity types, 4 relations) and inherently low inter-annotator agreement, which underscores the real-world complexity of this task. Our experiments systematically evaluated a small, open-weight LLM (Llama-3-8B) using few-shot prompting and parameter-efficient fine-tuning, comparing its performance against off-the-shelf biomedical IE systems across relevant span detection, named entity recognition, and relation extraction.

Key findings reveal that both few-shot prompting (with only 10 examples) and fine-tuned LLM approaches significantly outperform the specialized off-the-shelf systems, demonstrating the adaptability of LLMs even with minimal task-specific data. Despite this, absolute performance, particularly for complex relations and fine-grained entity distinctions, remains modest. This reflects the extreme low-resource nature of the study, the high diversity and skewness in annotations, and the inherent difficulty of the task, representative of clinical tasks with highly specific information needs.^37^ Performance was generally better for more frequent and clearly defined categories, such as for relevant span detection and extraction of `unlikely-rel` relations. Our error analysis highlighted persistent challenges including precise boundary detection, entity specificity, interpretation of negation and modality cues, and handling complex hierarchical anatomical references. Future work on adapting LLMs to the complex requirements of fine-grained annotation of clinical texts is required to establish more reliable solutions. Methods integrating structured knowledge such as expert ontologies or knowledge graphs with language models may be promising in this context.^38^

## Data Availability

All data produced in the present work are contained in the manuscript.

https://doi.org/10.17632/6kxnvb7yyg.1

## 6. Ethics

Ethics approval for this project was obtained from Melbourne Health Human Research Ethics Committee, approval number HREC/17/MH/308, approval date: 24 October 2017. Identifying details were removed from reports prior to annotation, including date and location of studies, names of patients and reporting radiologists, and if needed, names of any specific clinicians mentioned in the report (e.g., “I urgently notified Dr. XXX via telephone at 03:00am regarding these results”). Unfortunately, ethics approval has not been granted for public release of the data to date.

## 7. Acknowledgements

The authors would like to thank Professor Patrick Kwan and Professor Terence O’Brien of Monash University, who provided guidance on the context of Epilepsy diagnosis and documentation. THT and KV acknowledge funding from the Australian Research Council grant IC170100030.

## 8. Appendices

Appendices are available on Mendeley Data^39^ at http://dx.doi.org/10.17632/6kxnvb7yyg.1.

## Appendix A. Annotation scheme (Entities)

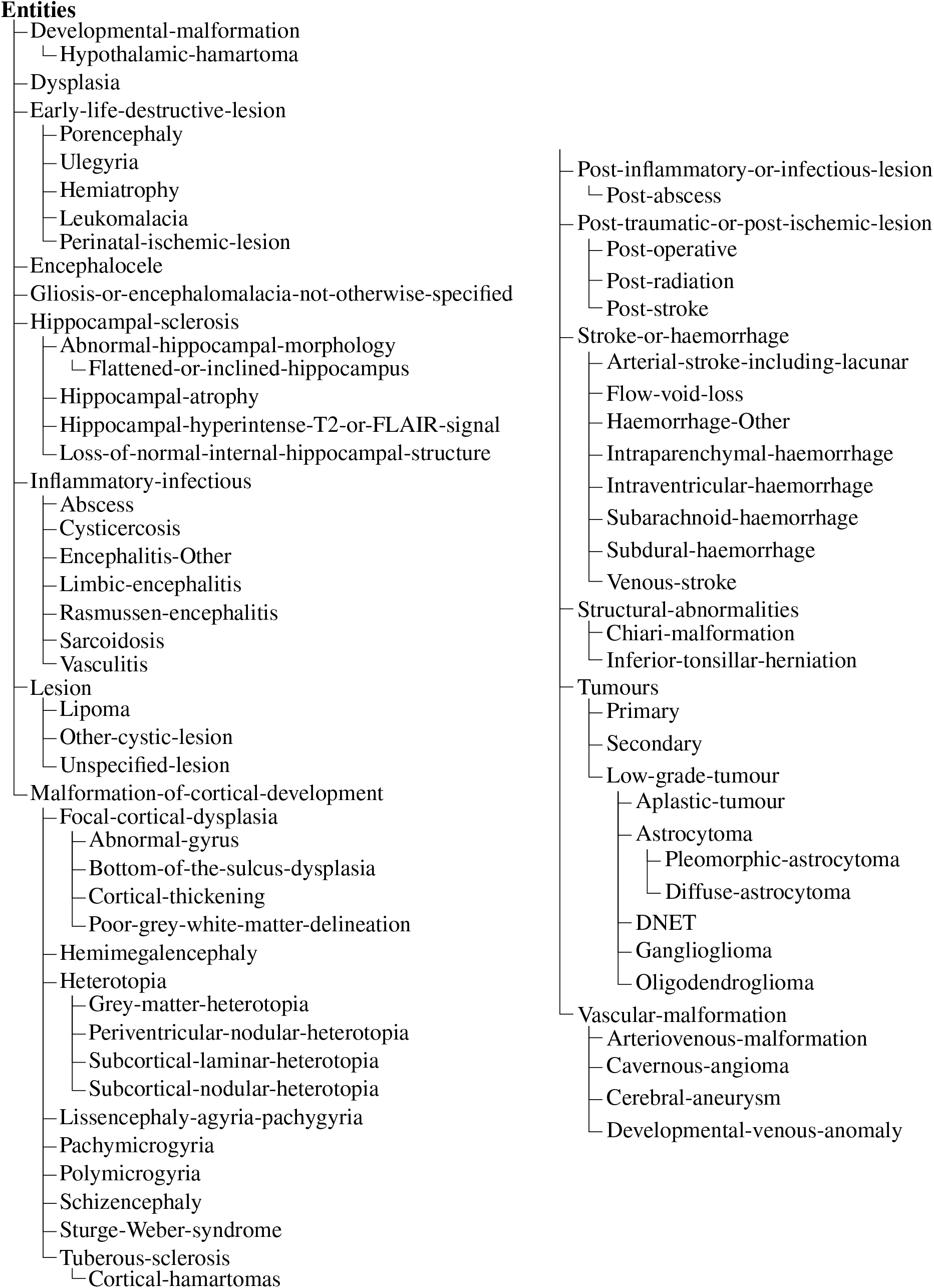

## Appendix B. Annotation scheme (Location Terms, Other Features, Hedging cues)

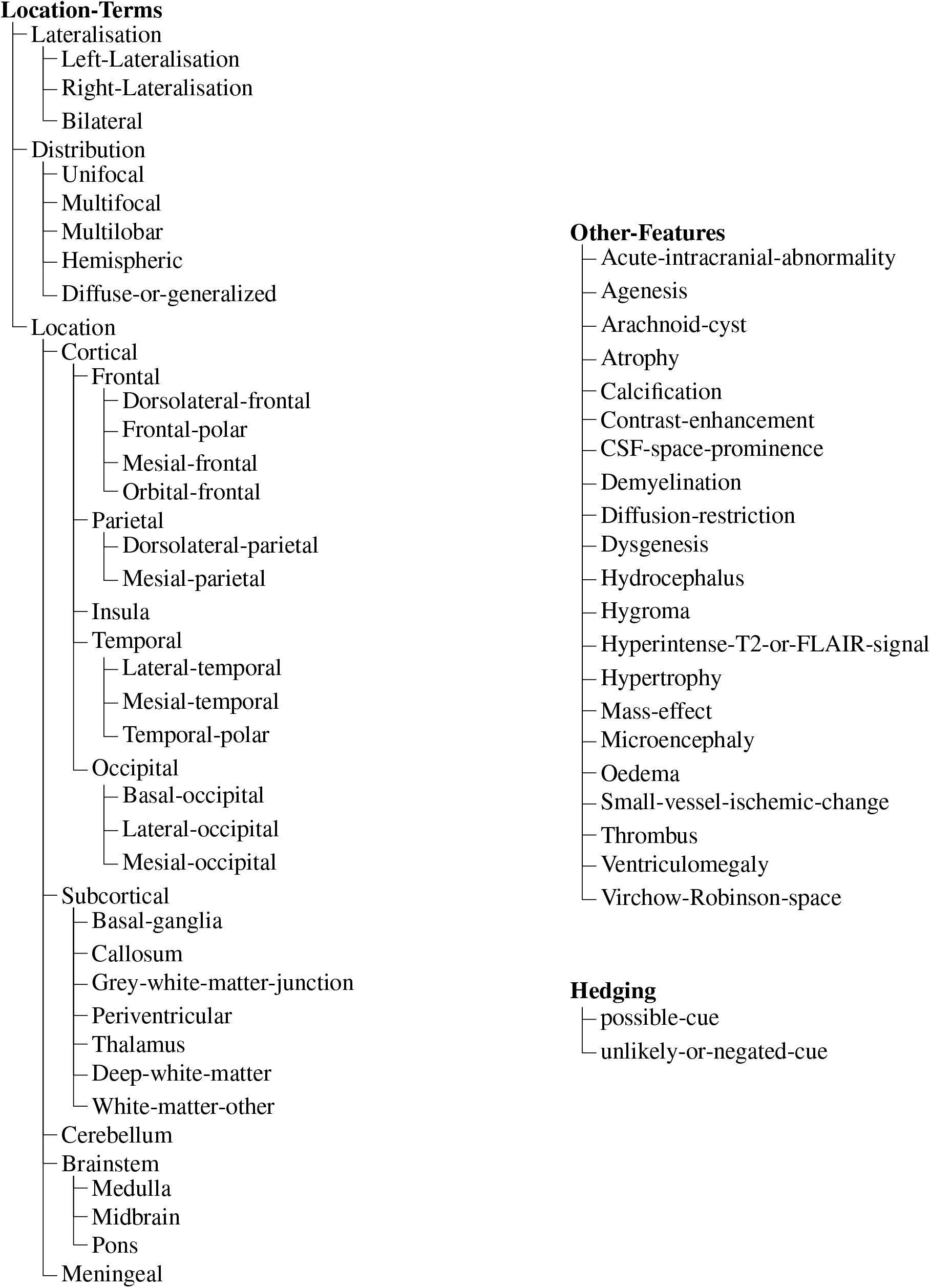

## Appendix C. Annotation scheme (Relations)

| Relation | Arguments | Description |
| --- | --- | --- |
| **possible-rel** | Arg1: possible-cue<br>Arg2: <LESION>\|<FEATURE> | Links possibility/uncertainty expressions to medical entities or features. Captures hedged or uncertain statements about findings. |
| **unlikely-rel** | Arg1: unlikely-or-negated-cue<br>Arg2: <LESION>\|<FEATURE> | Links negation/denial expressions to medical entities or features. Captures negative or ruled-out findings. |
| **location-rel** | Arg1: <LOCATION><br>Arg2: <LESION>\|<FEATURE> | Associates anatomical locations with medical entities or features. Specifies where findings are located anatomically. |
| **feature-rel** | Arg1: <FEATURE><br>Arg2: <LESION> | Links descriptive features to medical entities. Describes characteristics or properties of lesions. |

## Appendix D. Prompt details

### D.1. Relevant Span detection

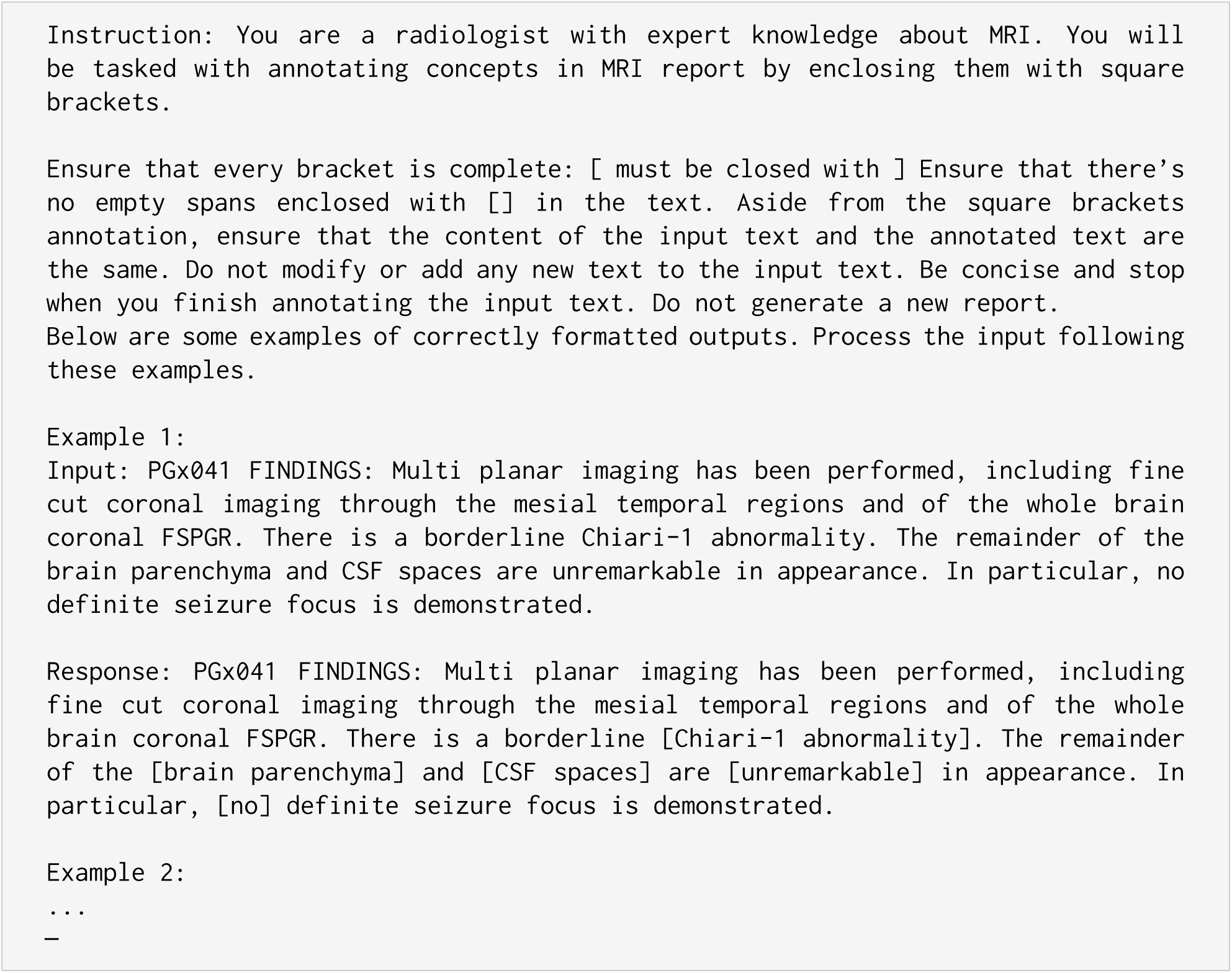

### D.2. Entity extraction

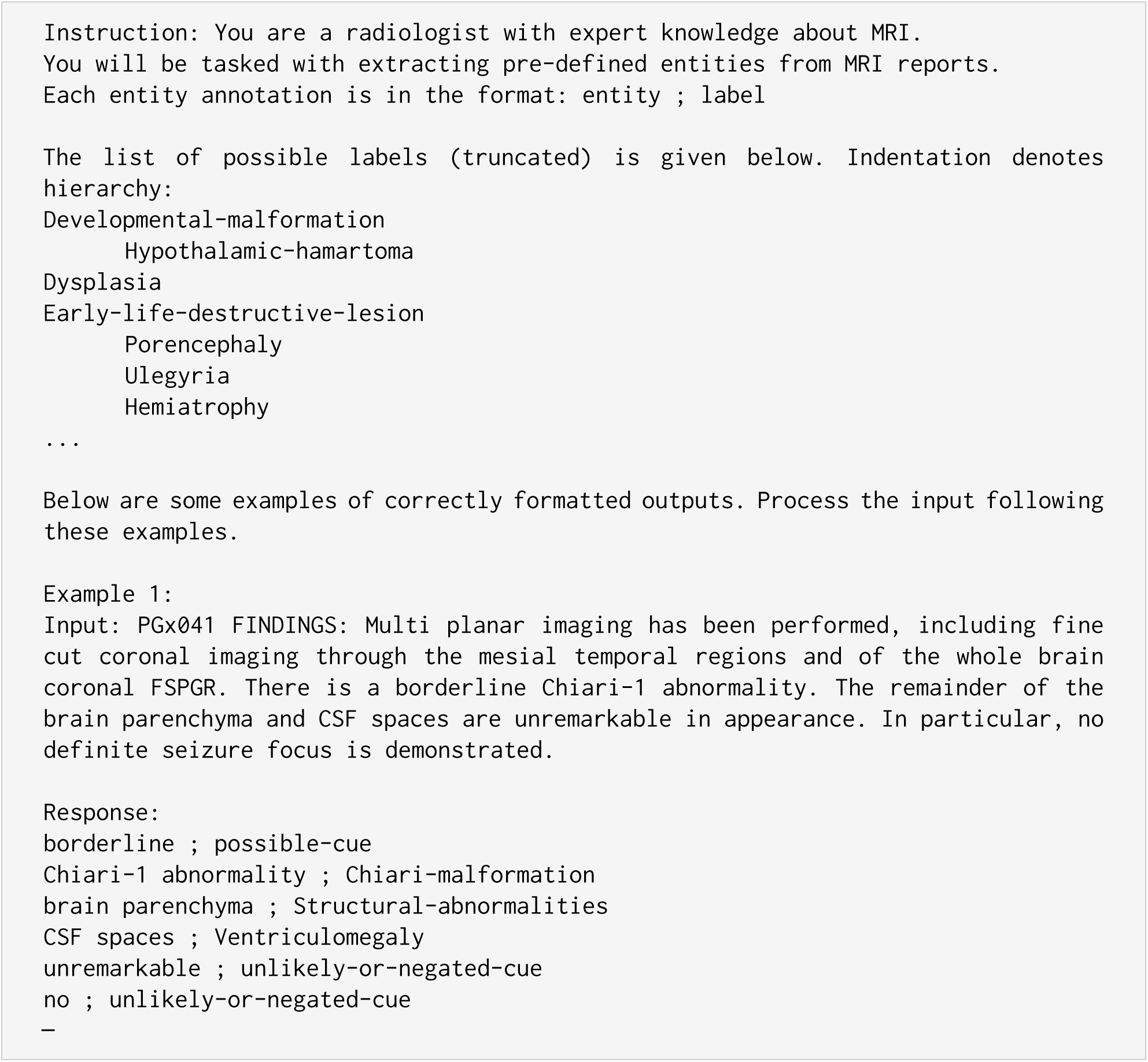

### D.3. Relation extraction

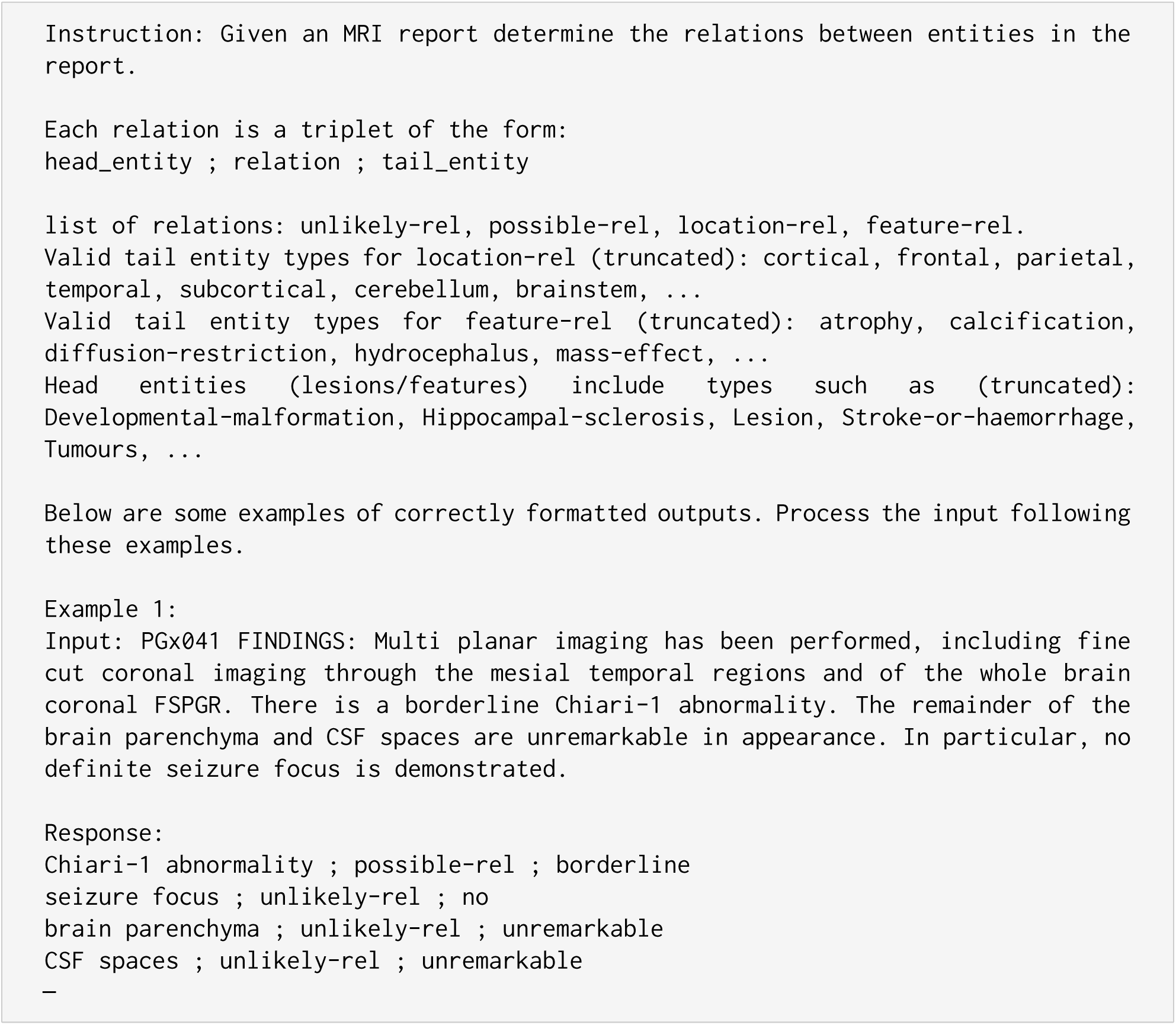

## Appendix E. Fine-tuning details

**Table E1.** Fine-tuning Configuration and Hyperparameters. Training is performed separately for each dataset level (inline, listing_coarse, listing, yaml) with 5 different random seeds.

| Parameter | Value |
| --- | --- |
| Base Model | Meta-Llama-3.1-8B-Instruct |
| Training Method | LoRA (Low-Rank Adaptation) |
| *LoRA Configuration* |  |
| Rank (r) | 16 |
| Alpha | 16 |
| Target Modules | q_proj, k_proj, v_proj, o_proj,<br>gate_proj, up_proj, down_proj |
| Dropout | 0 |
| Bias | none |
| *Training Parameters* |  |
| Batch Size | 2 |
| Gradient Accumulation Steps | 4 |
| Effective Batch Size | 8 |
| Learning Rate | 2e-4 |
| Weight Decay | 0.01 |
| Warmup Steps | 20 |
| Max Steps | 200 |
| LR Scheduler | linear |
| Optimiser | AdamW (8-bit) |
| *Model Configuration* |  |
| Max Sequence Length | 2048 |
| Mixed Precision | BF16 (if supported) or FP16 |
| Gradient Checkpointing | enabled (unsloth) |

## Footnotes

a http://www.commondataelements.ninds.nih.gov/

